# Genome-to-genome analysis reveals associations between human and mycobacterial genetic variation in tuberculosis patients from Tanzania

**DOI:** 10.1101/2023.05.11.23289848

**Authors:** Zhi Ming Xu, Michaela Zwyer, Daniela Brites, Hellen Hiza, Mohamed Sasamalo, Miriam Reinhard, Anna Doetsch, Sonia Borrell, Olivier Naret, Sina Rüeger, Dylan Lawless, Faima Isihaka, Hosiana Temba, Thomas Maroa, Rastard Naftari, Christian Beisel, Jerry Hella, Klaus Reither, Damien Portevin, Sebastien Gagneux, Jacques Fellay

**Author notes:** **Corresponding Author:** Jacques Fellay, School of Life Sciences, École Polytechnique Fédérale de Lausanne, 1015, Lausanne, Switzerland.

## Abstract

The risk and prognosis of tuberculosis (TB) are affected by both human and bacterial genetic factors. To identify interacting human and bacterial genetic loci, we leveraged paired human and *Mycobacterium tuberculosis* (*M*.*tb*) genomic data from 1000 Tanzanian TB patients. Through a genome-to-genome approach, we identified two pairs of human and *M*.*tb* genetic variants that are significantly associated. One of the human genetic variants maps to the intron of *PRDM15*, a gene involved in apoptosis regulation. The other human variant maps to an intergenic region close to *TIMM21* and *FBXO15*. In addition, we observed that a group of linked *M*.*tb* epitope variants were significantly associated with HLA-DRB1 variation. This suggests that even though epitope variation is rare in *M*.*tb* in general, specific epitopes might still be under immune selective pressure. Overall, our study pinpoints sites of genomic conflicts between humans and *M*.*tb*, suggesting bacterial escape from host selection pressure.

## Introduction

Tuberculosis (TB) is an infectious disease mainly caused by *Mycobacterium tuberculosis* (*M*.*tb*). The infection primarily affects the lungs, although extrapulmonary TB manifests in around 20-25% of cases^1^. TB continues to pose a significant public health challenge, particularly in low- and middle-income countries. Approximately 10 million people develop active TB yearly, resulting in 1.6 million deaths^2^. Except in 2022 after the COVID-19 pandemic, TB remains the leading cause of death from an infectious disease worldwide^2,3^.

Exposure to *M*.*tb* can lead to a wide range of clinical manifestations^4,5^. Most infected individuals develop latent TB, where quiescent infection can heal or persist but remain asymptomatic^6^. However, latent TB might progress into active TB disease in 5%-15% of cases^7^, where symptoms such as cough, fever, or weight loss develop. Several factors modulate the risk of developing active TB, including HIV co-infection, which increases the risk by almost 20-fold^8^. Additional risk factors include malnutrition, alcohol abuse, smoking, and other comorbidities such as diabetes^9–11^.

Both human and bacterial genetic factors modulate the risk of developing active TB and its prognosis. Early twin studies have suggested that TB susceptibility has a heritable component^12,13^. In the past decade, genome-wide association studies (GWAS) have identified many genetic loci involved in TB susceptibility^14–23^. However, a significant limitation of GWAS of TB is that associations failed to replicate across studies in different population^24^. This might be explained in part by a failure to consider bacterial genetic variation. Human-adapted *M*.*tb* strains are classified into nine lineages (L1-L9)^25^, which can be further classified into sublineages according to phylogenetic markers^26^. Despite *M*.*tb* strains being highly clonal^27^, different lineages show some difference in pathogenicity^28^. The prevalence of some lineages is also highly geographically structured^29^. While this could be partially due to genetic drift or reflect migration patterns, another possibility is that this reflects adaptation of lineages to human populations with particular genetic backgrounds^30,31^.

A hypothesis-free method to identify interacting human and pathogen genetic loci is to jointly analyze paired pathogen and human genomes isolated from patients who have developed the respective disease. Specifically, a genome-to-genome (G2G) approach could be used, where associations between all pairs of host and pathogen genetic variants are tested^32^. Significant associations identified under this framework reflect combinations of host and pathogen genotypes that jointly modulate the risk of developing an infectious disease upon exposure to the pathogen. In addition, this approach can also identify pathogen loci that have undergone intra-host selection driven by specific host genetic variants.

In this study, we leveraged paired *M*.*tb* and human genomic data in a cohort of 1000 active TB patients recruited in Dar es Salaam, Tanzania. Using a G2G approach, we identified significant associations between humans and *M*.*tb* genetic variants, indicating sites of genomic conflict.

## Results

### Study description

This study was based on a cohort of 1000 active TB patients recruited in Dar es Salaam, Tanzania (TB-DAR) for which we generated both *M*.*tb* and human genomic data. Table 1 summarizes the demographic and clinical characteristics of the cohort. In brief, the cohort consisted of 70% males and 30% females, with a mean age of 34. 16% of patients were co-infected with HIV. Patients were infected with *M*.*tb* lineage L3 (43.1%), L4 (33.2%), L1 (15.5%) and L2 (8.2%).

**Table 1.**
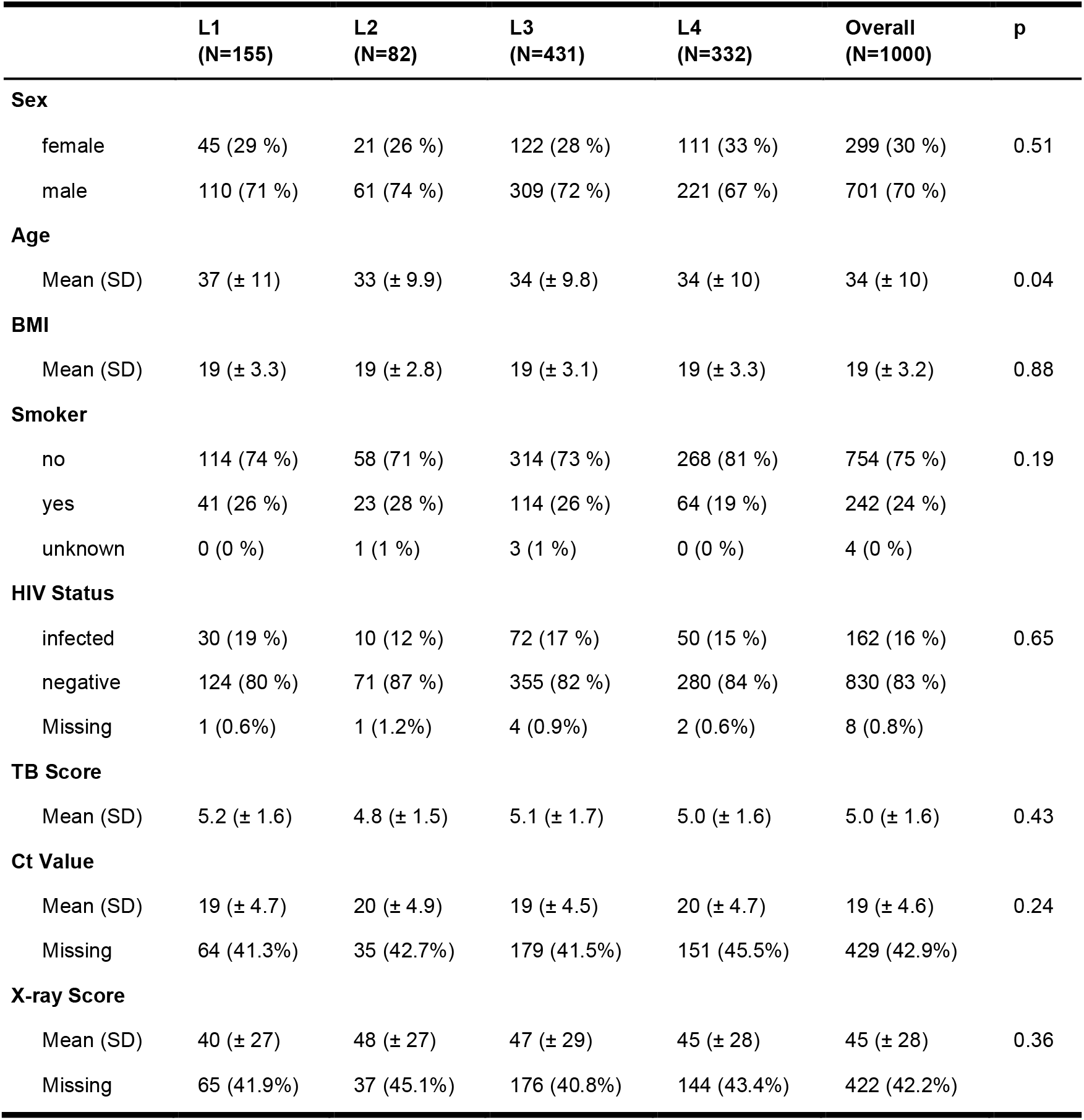
Demographic and clinical characteristics of the cohort. L1-L4 indicates bacterial lineages. P value based on ANOVA test for quantitative variables and chi-squared test for discrete variables.

|  | L1<br>(N=155) | L2<br>(N=82) | L3<br>(N=431) | L4<br>(N=332) | Overall<br>(N=1000) | p |
| --- | --- | --- | --- | --- | --- | --- |
| Sex |  |  |  |  |  |  |
| female | 45 (29 %) | 21 (26 %) | 122 (28 %) | 111 (33 %) | 299 (30 %) | 0.51 |
| male | 110 (71 %) | 61 (74 %) | 309 (72 %) | 221 (67 %) | 701 (70 %) |  |
| Age |  |  |  |  |  |  |
| Mean (SD) | 37 (± 11) | 33 (± 9.9) | 34 (± 9.8) | 34 (± 10) | 34 (± 10) | 0.04 |
| BMI |  |  |  |  |  |  |
| Mean (SD) | 19 (± 3.3) | 19 (± 2.8) | 19 (± 3.1) | 19 (± 3.3) | 19 (± 3.2) | 0.88 |
| Smoker |  |  |  |  |  |  |
| no | 114 (74 %) | 58 (71 %) | 314 (73 %) | 268 (81 %) | 754 (75 %) | 0.19 |
| yes | 41 (26 %) | 23 (28 %) | 114 (26 %) | 64 (19 %) | 242 (24 %) |  |
| unknown | 0 (0 %) | 1 (1 %) | 3 (1 %) | 0 (0 %) | 4 (0 %) |  |
| HIV Status |  |  |  |  |  |  |
| infected | 30 (19 %) | 10 (12 %) | 72 (17 %) | 50 (15 %) | 162 (16 %) | 0.65 |
| negative | 124 (80 %) | 71 (87 %) | 355 (82 %) | 280 (84 %) | 830 (83 %) |  |
| Missing | 1 (0.6%) | 1 (1.2%) | 4 (0.9%) | 2 (0.6%) | 8 (0.8%) |  |
| TB Score |  |  |  |  |  |  |
| Mean (SD) | 5.2 (± 1.6) | 4.8 (± 1.5) | 5.1 (± 1.7) | 5.0 (± 1.6) | 5.0 (± 1.6) | 0.43 |
| Ct Value |  |  |  |  |  |  |
| Mean (SD) | 19 (± 4.7) | 20 (± 4.9) | 19 (± 4.5) | 20 (± 4.7) | 19 (± 4.6) | 0.24 |
| Missing | 64 (41.3%) | 35 (42.7%) | 179 (41.5%) | 151 (45.5%) | 429 (42.9%) |  |
| X-ray Score |  |  |  |  |  |  |
| Mean (SD) | 40 (± 27) | 48 (± 27) | 47 (± 29) | 45 (± 28) | 45 (± 28) | 0.36 |
| Missing | 65 (41.9%) | 37 (45.1%) | 176 (40.8%) | 144 (43.4%) | 422 (42.2%) |  |

As part of the study, we collected clinical measures including TB score, X-ray score, and GeneXpert Ct-value. TB score is a symptoms-based score. X-ray score corresponds to lung damage severity based on chest x-ray images. GeneXpert Ct-value is based on RT-PCR, and is inversely correlated with sputum bacterial load. We observed that patients infected with different lineages did not present significantly different clinical characteristics (ANOVA Test; TB Score: p = 0.43, X-ray score: p = 0.24, and Ct-value: p = 0.36).

### Associations between *M*.*tb* and human genetic variants

We tested for associations between all human and pathogen variants. Based on statistical power calculations (Supplementary Figure 1), we restricted our analysis to 1538 common *M*.*tb* amino-acid variants with minor-allele frequency (MAF) > 0.015 and 6,603,291 common human variants with MAF > 0.05. We corrected for population stratification using the top three human principal components (PCs). Genomic inflation factors (λ) were below 1, indicating the absence of inflation of test statistics due to stratification (Supplementary Figure 2).

Considering that a large proportion of *M*.*tb* variants were perfectly correlated (Supplementary Figure 3), many association tests were not independent (Supplementary Figure 4). Therefore, we derived a multiple-testing corrected p-value threshold (p<1.02e-10) based on the number of independent tests and the genome-wide significant threshold of 5e-8 for the human genome (See Methods).

We identified two significant G2G associations (Figure 1). The first association was between the *M*.*tb* variant Rv2348c I101M and the human single nucleotide polymorphism (SNP) rs12151990 (p = 4.7e-11, OR = 5.6). rs12151990 is an intronic SNP in *PRDM15* located on chromosome 21 (Supplementary Figure 5A). Colocalization of GWAS and *PRDM15* eQTL signals suggested shared causal variants in some tissues (Supplementary Figure 6A), albeit not in the lung. According to gnomAD, rs12151990 is common (MAF > 0.05) in all populations except South Asians. On the bacterial side, Rv2348c I101M was prevalent in lineage 4 (MAF = 0.051) but also found in L1 (MAF = 0.007) in our cohort. Within L4, the variant belonged to a subclade within sublineage L4.3 but also displayed homoplasy on three occasions which could indicate positive selection (Figure 2). Indeed, when looking for this variant in a global reference set of L1-L4 *M*.*tb* genomes (n = 11,818) compiled by Zwyer et al^33^, we saw that the mutation had arisen independently multiple times within each lineage, suggesting that it is under positive selection. Using PAML, we formally tested for positive selection on a subset of 500 randomly selected strains from the reference set and on a randomly selected subset of 300 genomes for each lineage separately. We found that Rv2348c was indeed under positive selection in L1 and L4 (p<0.0001), but not in L2 and L3 (p=0.48 and 0.40, respectively). Moreover, it corresponds to the only codon within Rv2348c that was identified to be under positive selection in L1 and L4 (posterior probability > 99%).

**Figure 1.**
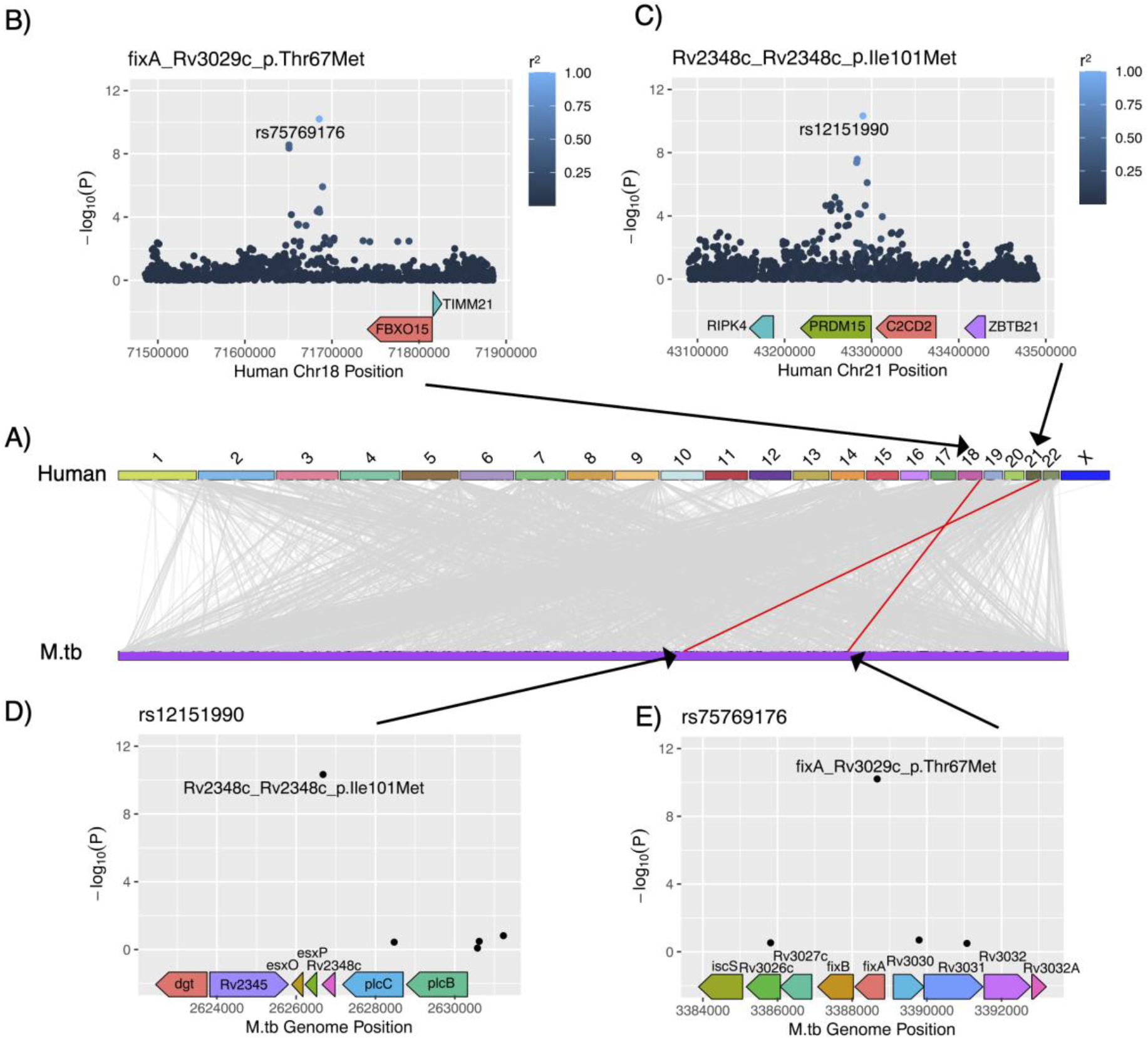
Summary of genome-to-genome (G2G) analysis results. **A)** Associations between human and *M*.*tb* variants, with upper half of the figure refers to human (GRCh37) chromosome coordinates and bottom half refers to *M*.*tb* (H37Rv) nucleotide positions. Grey lines indicate genome-wide significant (p < 5e-8) G2G associations and red lines indicates significant associations after Bonferroni correction (p < 1.02e-10) **B)** Manhattan plot of human genetic loci referring to the association between FixA T67M and rs75769176 **C)** Manhattan plot of human genetic loci referring to the association between Rv2348c I101M and rs12151990 **D)** Manhattan plot of bacterial genetic loci referring to the association between Rv2348c I101M and rs12151990 **E)** Manhattan plot of bacterial genetic loci referring to the association between FixA T67M and rs75769176

**Figure 2.**
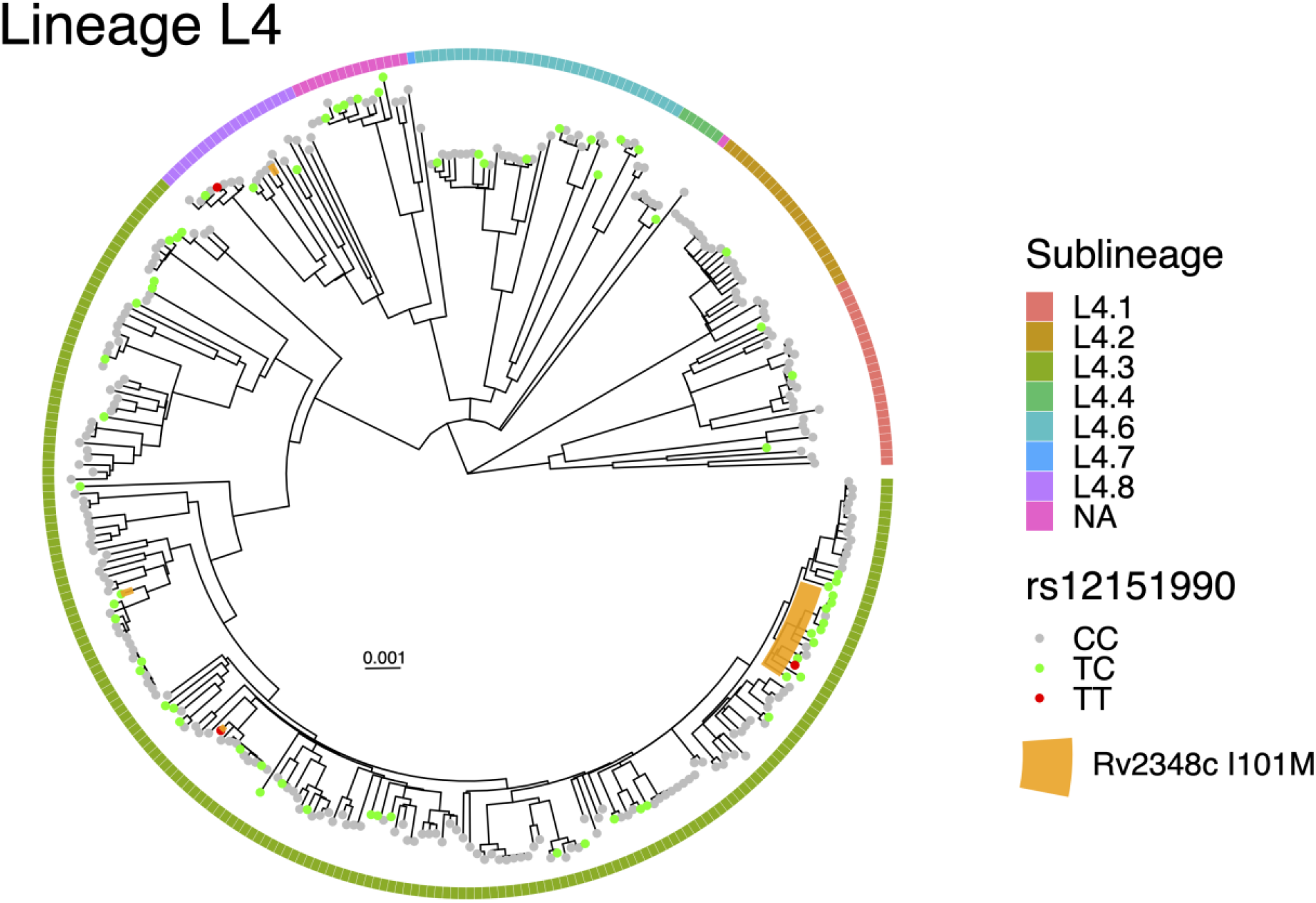
Phylogenetic tree of *M*.*tb* strains in Lineage 4. Colors of outermost squares refer to sublineages. Colors of circles on tree tips refer to whether the infected human carries the rs12151990 variant. Orange shade represents the clade (or in 3 instances, single strains) of *M*.*tb* that carry the Rv2348c I101M variant. The tree was rooted with a *M. canetti* strain as an outgroup. Scale bar represents substitutions per polymorphic site.

The second association was between the *M*.*tb* variant FixA T67M and the human SNP rs75769176 (p = 6.3e-11, OR = 6.7). rs75769176 maps to an intergenic region close to *FBXO15* and *TIMM21* on chromosome 18 (Supplementary Figure 5B). The GWAS signal did not colocalize with eQTLs signals from either gene (Supplementary Figure 6B and 6C). According to gnomAD, rs75769176 is specific to African populations (MAF = 0.08). On the bacterial side, FixA T67M was found exclusively in lineage L3 (MAF = 0.053) and belonged to a subclade within sublineage L3.1 in our cohort. When investigating the global reference dataset, we still found the variant exclusively in sublineage L3.1.

In an attempt to identify potential confounders, we tested whether the two *M*.*tb* variants were associated with any patient characteristics (Supplementary Table 1 and Supplementary Table 2). The results indicated that no clinical measures were significantly associated with the *M*.*tb* variants. Importantly, HIV status was also not significantly associated. We observed that FixA T67M was associated with the second human genetic PC. However, this PC was already included as a covariate in the G2G study to prevent spurious associations driven by population stratification.

We next tested whether the G2G-associated human and bacterial variants were associated with any clinical measures of TB (Supplementary Table 3). On the bacterial side, we did not identify variants significantly associated with any clinical measure. On the human side, the minor allele at rs75769176 was negatively associated with TB score (p = 0.04, log(OR) = -0.34), indicating an association with less severe disease.

Finally, we investigated whether human variants previously found to be associated with susceptibility to developing active TB were also associated with *M*.*tb* variation in our study (Supplementary Table 4). For each human variant, we extracted the most significant G2G-associated *M*.*tb* variant. For human variants that were rare within our cohort, we tested all common variants within ±5kb and extracted the top association. The strongest association we identified was between rs4240897 and Rv2963 K165fs (p = 9.5e-5, OR = 1.4). We also repeated the same analysis but with human variants identified in previous host-pathogen studies to be associated with *M*.*tb* lineages or clades (Supplementary Table 5). The strongest association we identified was between rs146731249 (a proxy for rs17235409) and HsdM K483E (p = 5.0e-6, OR = 4.8).

### Interactions between *M*.*tb* and human pathways

To identify whether G2G associations were enriched for certain molecular functions, we leveraged human and *M*.*tb* pathways from existing gene regulatory network databases. Human pathways were extracted from the Molecular Signatures Database (MSigDB). *M*.*tb* pathways were either extracted from a co-regulatory network database (MTB Network Portal) or constructed from stringDB. To construct pathway-to-pathway edges between all pairs of human and *M*.*tb* pathways, we mapped variants to genes and binarized G2G associations by applying a lenient threshold (FDR < 0.15). We then calculated the density of pathway-to-pathway edges between all pairs.

To search for enriched pairs of human and *M*.*tb* pathways, we compared the true densities to those found based on permuted datasets. The pair of pathways with the highest enrichment of G2G associations had a density of 0.11 (3 edges / 28 possible edges). However, the enrichment was not statistically significant (p=0.07) compared to the distribution derived from permutations (Supplementary Figure 7).

### Associations between variants in *M*.*tb* T cell epitopes and human HLA variation

Escape mutations in T cell epitopes are commonly observed in pathogens causing chronic infections to avoid HLA-driven cellular immunity. To search for potential HLA-induced *M*.*tb* variants, we extracted experimentally validated T-cell epitopes from the Immune Epitope Database (IEDB) and retained those that were polymorphic within our cohort (frequency > 0.015). Fifteen *M*.*tb* variants overlapped with at least one epitope: 9 epitope variants were independent, while 4 variants were perfectly correlated with each other and thus grouped into a single variant set (Supplementary Figure 8). On the human side, we imputed HLA amino acid variants and HLA 4-digit alleles using a trans-ethnic reference panel TopMed.

For each *M*.*tb* epitope variant, we tested whether it was associated with any human HLA allele or amino acid variant. To correct for multiple testing, we applied a Bonferroni corrected significance threshold of 1.15e-5 based on the number of epitope variants, the number of human HLA amino acid positions or alleles, and an alpha level of 0.05. A significant association was identified between an epitope variant set (EspK L39W, EsxB E68K, Mpt70 A21T, RimJ R72L) and HLA-DRB1 H96E (p = 7.3e-06, OR = 6.31). The variant EsxB E68K maps to epitopes that have been experimentally validated to be restricted by various HLA-DRB1 alleles (Supplementary Table 6).

Finally, we extracted the top association for every *M*.*tb* epitope variant and compared that against the top associations for *M*.*tb* variants that are not part of any annotated T cell epitope. We observed that the association between the epitope variant set (EspK L39W, EsxB E68K, Mpt70 A21T, RimJ R72L) and HLA-DRB1 H96E was stronger than any other associations (Figure 4). Furthermore, associations between epitope variants and HLA amino acids were on average stronger than associations between variants that were not part of any annotated T cell epitope and HLA amino acids (p = 0.015) (Figure 4). A similar trend was observed for HLA 4-digit alleles (p = 0.003).

**Figure 3.**
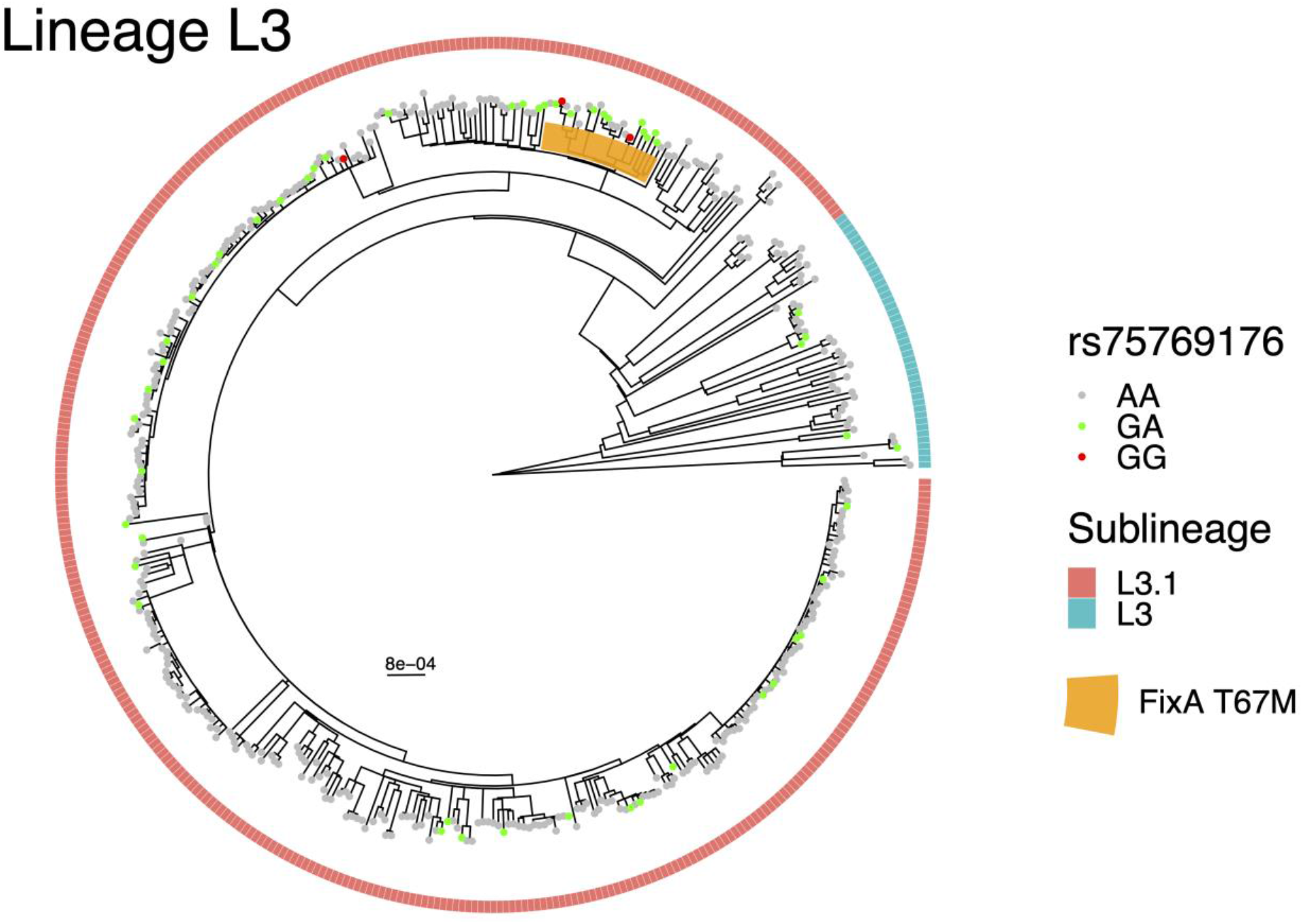
Phylogenetic tree of *M*.*tb* strains in Lineage L3. Colors of outermost squares refer to sublineages. Colors of circles on tree tips refer to whether the infected human carries the rs75769176 variant. Orange shade represents the clade of *M*.*tb* that carry the FixA T67M variant. The tree was rooted with a *M. canetti* strain as an outgroup. Scale bar represents substitutions per polymorphic site.

**Figure 4.**
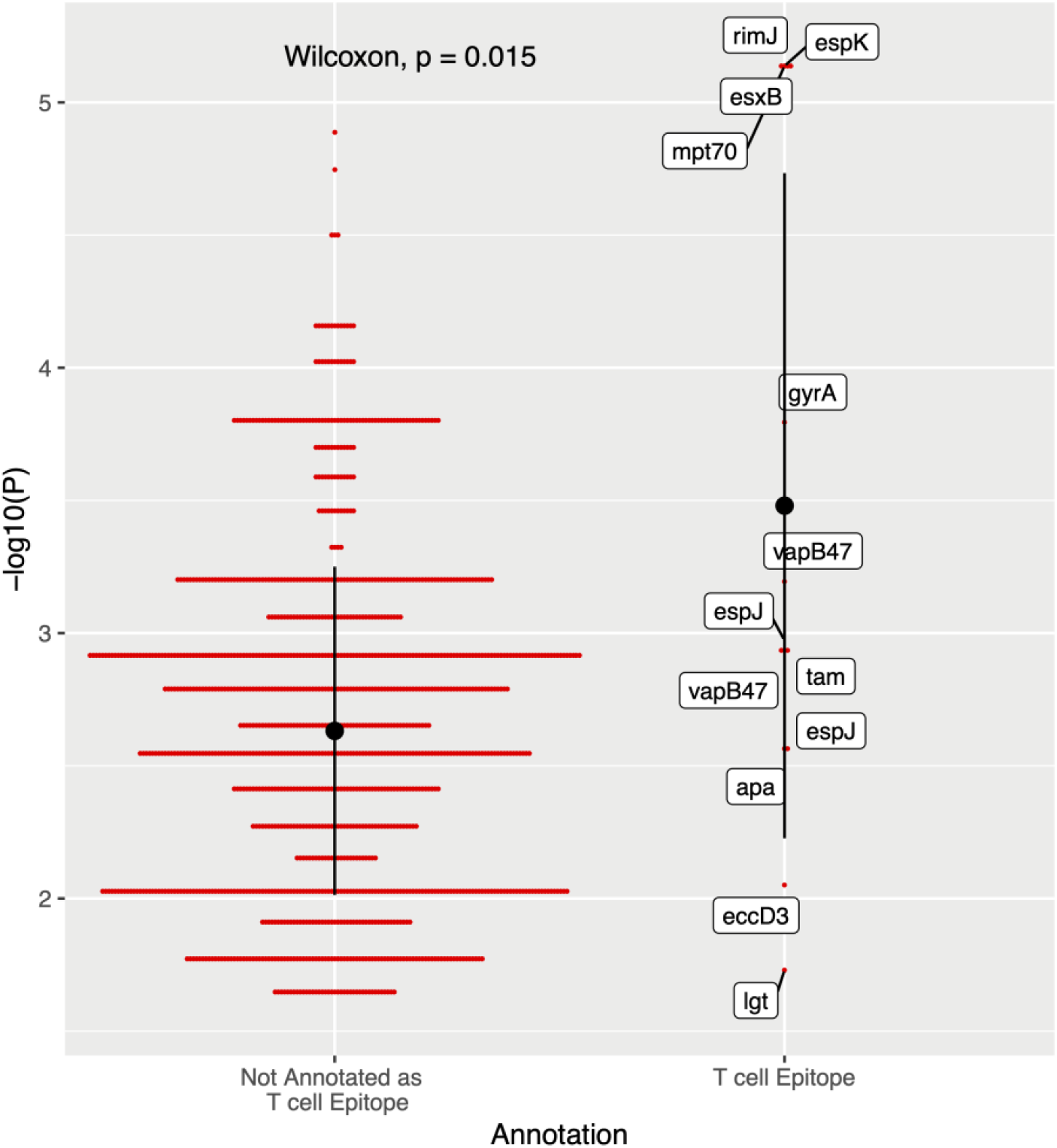
Associations between *M*.*tb* variants and HLA amino acid variants. For each *M*.*tb* variant, only the top association is displayed. Not annotated as T cell epitope: *M*.*tb* variants that are not part of any annotated T cell epitope; T cell epitope: *M*.*tb* variants that map to known T cell epitopes based on the IEDB database. Black dots and bars indicate mean and standard deviation within each annotation category.

## Discussion

We describe the first TB genome-to-genome (G2G) study conducted in an African population. We identified two pairs of associated genetic loci that were not identified in previous G2G studies or GxG interaction studies^34–38^. In addition, we show that although variations in *M*.*tb* epitopes are rare, certain *M*.*tb* epitope variants are associated with human HLA variation, a sign of probable immune selective pressure.

The first G2G association we identified was between the *M*.*tb* variant Rv2348c I101M and the human SNP rs12151990. rs12151990 is an intronic variant that affects the mRNA expression of *PRDM15* in some tissues. A previous study has shown that *PRDM15* is co-expressed with *KCNJ15* - a pro-apoptotic gene that is differentially acetylated when comparing blood monocytes and granulocytes derived from *M*.*tb* infected patients and healthy controls^39^. This suggests that *PRDM15* could be involved in regulation of apoptosis, an important defense mechanism that human macrophages employ for *M*.*tb* clearance^40,41^. The function of Rv2348c is unknown, other than that it is an antigen that stimulates T cell-mediated responses^42,43^. Since the induction of apoptosis in infected macrophages facilitates the presentation of antigens by dendritic cells and subsequent activation of T cell-mediated responses^44^, it is possible that Rv2348c and *PRDM15* jointly determine disease prognosis due to such a process. Alternatively, it is possible that Rv2348c might be directly involved in mechanisms that M.tb employs to regulate host cell apoptosis^45^.

The second G2G association we identified was between the *M*.*tb* variant FixA T67M and the human SNP rs75769176. rs75769176 maps to an intergenic region and is close to both *FBXO15* and *TIMM21*. Since colocalization analysis did not reveal any shared GWAS-eQTL signals for the two genes, it is unclear whether the SNP impacts regulation of gene expression. It is important to note that such analysis has limited power given that large tissue-specific eQTL studies were only available for European populations, and it has been shown that a large proportion of eQTLs can be population-specific^46^. According to Reactome, *FBXO15* belongs to the “Class I MHC mediated antigen processing and presentation” pathway suggesting its involvement in regulating adaptive immunity. *TIMM21* belongs to the “Mitochondrial protein import” pathway. Again, the exact mechanism for which FixA is involved in the interaction is unclear, given that limited information suggests its involvement in *M*.*tb* metabolism and respiration. Nevertheless, the association of rs75769176 with TB severity (TB score) in our cohort suggests that the variant plays a role in TB pathogenesis.

Based on phylogenetics, we believe the two G2G-associated *M*.*tb* variants may have evolved under different scenarios. FixA T67 did not display homoplasy but strictly belonged to a clade within L3.1. The G2G-associated human SNP (rs75769176) is associated with lower severity (TB score). This could correspond to the scenario where individuals who carry the protective human variant are more susceptible to developing active TB upon exposure to *M*.*tb* strains that carry the FixA T67 variant compared to *M*.*tb* strains that do not. Similarly, most strains with Rv2348c I101M belonged to a clade within L4.3. However, Rv2348c I101M also displayed homoplasy, suggesting that in some cases the mutation might have undergone intra-host selection during infection similar to *de novo* drug resistance mutations^47^. Specifically, Rv2348c I101M might confer a fitness advantage for the bacteria within carriers of rs12151990 and have outcompeted the ancestral strain that the carriers were originally infected with.

Corresponding to previous findings, we observed that *M*.*tb* T cell epitopes are hyperconserved^48^: in our study, only 15 out of 1,538 common *M*.*tb* variants overlapped with known T cell epitopes. This suggests that epitope variation may not be the primary mechanism that *M*.*tb* employs to evade T cell response and that alternative mechanisms exist^49^. For example, one proposed mechanism is that *M*.*tb* secretes “decoy” antigens that are non-essential for bacterial survival during the initial stages of infection. As a result, T cells would be primed against these antigens and subsequently fail to recognize the pathogen during the later stages of infection once the “decoy” antigens are not expressed^50^. For these antigens, variation provides no selective advantage, given that the goal is to subvert rather than evade. Nevertheless, a small subset of epitopes does display variation, and it is hypothesized that these epitopes may be in antigens that employ a different evasion mechanism from those in hyper-conserved antigens^51^. A recent study has identified region-specific variation in some *M*.*tb* epitopes, suggesting they may be under selective pressure induced by human HLA variation^52^. Our results further support this hypothesis by providing direct genetic evidence on the role of human HLA-DRB1 in inducing *M*.*tb* epitope variations. Importantly, these epitopes could potentially be targeted by future T cell vaccines, given that they are likely under immune selective pressure and thus more likely to be important for host control of the bacteria.

The first limitation of applying a G2G approach to study host-pathogen interactions in TB is that *M*.*tb* is highly clonal and many *M*.*tb* variants are strongly linked. For example, HLA-DRB1 H96E is associated with four epitope variants and other non-annotated variants. While some of the variants may be linked due to epistasis, many others may be linked simply due to evolutionary history. Therefore, further experimental studies are needed to elucidate the causal variants. The second limitation of our study is that only active TB cases were recruited. In the absence of a comparable analysis in subclinical or latent TB groups, it is not possible to know if the observed enrichment of certain combinations of host and pathogen variants (G2G associations) increases the risk of developing active TB or results from host selection pressure during active mycobacterial replication. A third limitation is that patients seek hospital care at different stages of the disease, rendering clinical outcome measurements less reliable. We tried to correct for this by adding cough duration as a covariate when comparing clinical data across patient groups. Nevertheless, given its self-reported nature, cough duration might not be a perfect proxy for disease length. Finally, given that the bacterial isolates were cultured prior to sequencing, the full intra-host diversity of bacterial populations may not be fully recapitulated. Certain strains may have growth advantages and thus may be overrepresented. This did not affect our analysis since we focused on the consensus sequence and ignored sites with intra-host diversity. Indeed, when considering variants that are fixed intra-host, sputum-based sequencing was found to be concordant with culture-based sequencing^53^. Nevertheless, direct deep-sequencing of sputum samples could be useful to explore the intra-host diversity of *M*.*tb*, especially in the context of low-frequency *de novo* mutations.

## Conclusion

Through a genome-to-genome (G2G) analysis of TB patients from Tanzania, we identified two pairs of associated genetic loci that might play a role in TB pathogenesis. The association between the *M*.*tb* variant Rv2348c I101M and the human SNP rs12151990 could be due to the interplay between *M*.*tb* and host apoptosis responses, although this requires further validation. The mechanism responsible for the association between the *M*.*tb* variant FixA T67M and the human SNP rs75769176 is unknown. However, the human SNP was associated with TB severity, indicating a potential impact on TB pathogenesis. In addition, we observed that a set of *M*.*tb* epitope variants (EspK L39W, EsxB E68K, Mpt70 A21T, RimJ R72L) were associated with HLA-DRB1 H96E. This shows that certain *M*.*tb* epitope mutations are under selective pressure induced by human HLA variation, confirming a role for T cell immunity in *M*.*tb* control. Our results highlight genomic loci involved in the host-pathogen battle during chronic infection with *M*.*tb*. A deeper understanding of the mechanisms at play could be instrumental in developing new therapeutic strategies against TB.

## Methods

### Recruitment and sample collection

This study included active TB patients (sputum smear-positive and GeneXpert-positive) recruited at the Temeke District Hospital in Dar es Salaam, Tanzania, as part of a prospective study that ran between November 2013 and June 2022 (TB-DAR cohort). Ethical approval for the TB-DAR cohort has been obtained from the Ethikkomission Nordwest-und Zentralschweiz (EKNZ UBE-15/42), the Ifakara Health Institute—Institutional Review Board Board (IHI/IRB/EXT/No: 24–2020) and the National Institute for Medical Research in Tanzania—Medical Research Coordinating Committee (NIMR/HQ/R.8c/Vol.I/1622). A written informed consent has been obtained from every patient who has been recruited into the TB-DAR cohort.

Clinical information including TB score, bacterial load (GeneXpert Ct-value), X-ray score, and HIV status was collected as part of the study. TB-score is a symptoms-based score (with a maximum of 12 points) adapted from Wejse et al^54^. A point was added for the presence of each of the following signs or symptoms: cough, hemoptysis, dyspnea, chest pain, night sweat, anemia, abnormal auscultation, body temperature above 37°C, BMI below 18, BMI below 16, mid-upper arm circumference (MUAC) below 220, MUAC below 200. Bacterial load was measured using the GeneXpert Ct-value from real-time polymerase chain reaction. X-ray score is based on the Ralph score^55^ and assigned by two independent radiologists for patients with high-quality chest X-ray images. Both Ct-value and X-ray score were transformed using rank-based inverse normal transformation^56^. Demographic information including age, sex, and self-reported smoking status was also collected. Patients recruited twice due to relapse or reinfection were excluded.

In total, we generated either human or bacterial genomic data for 1,906 patients. For 1,471 patients, bacterial genomic data were generated. For 1,444 patients, human genomic data were generated (98 based on WGS, 1,384 based on genotyping, and 30 based on both). This study focused on 1,000 patients where high-quality human and bacterial genomic data were both available after quality-based filtering.

### Human sequencing

WGS was performed at the Health 2030 Genome Center in Geneva, Switzerland. The Illumina NovaSeq 6000 sequencer was used, starting with 1 μg of whole blood genomic DNA. Illumina TruSeq DNA PCR-Free reagents were used for library preparation. The 150nt paired-end sequencing configuration was applied. An average coverage of 30X was achieved for each genome.

The BWA^57^ aligner (v0.7.17) was used to align sequencing reads to the GRCh38 (GCA 000001405.15) reference genome. Duplicate reads were marked using the markduplicates module of Picard^58^ (v2.8.14). For variant calling, GATK best practices (Germline short variant discovery) were followed. Base quality score recalibration was applied using the GATK. Variants were first called individually per sample. Then, samples with less than 5X coverage were excluded and variants were jointly called. A Quality Score Recalibration (VQSR) based filter was applied (truth sensitivity of 99.7 and excess heterozygosity of 54.69). Samples with a high rate of missing genotype calls (>50%) were also excluded.

### Human genotyping and imputation

Genotyping was performed by the iGE3 Genomics platform at the University of Geneva in Geneva, Switzerland. Illumina Infinium H3Africa genotyping microarrays (Version 2; https://chipinfo.h3abionet.org) with custom SNP add-ons (Tanzanian-specific SNPs, as described by Xu et al.^59^) were used. The Illumina GenomeStudio software (v2.0.5; https://support.illumina.com/array/array_software/genomestudio/downloads.html) was used to analyze raw microarray data. Low-quality or poorly clustered probes were excluded based on filters suggested by Illumina for human studies (Supplementary Table 7). Low-quality samples (call rate <0.97) were also excluded. Files were converted to PLINK format using the GenomeStudio PLINK Input Report Plug-in(v2.1.4) and to VCF using PLINK(v1.9)^60^.

Imputation was performed using two reference panels: 1) The African Genome Resources (AFGR) reference panel (https://www.apcdr.org/) and 2) An internal Tanzanian reference panel based on the 118 WGS samples from this study. For the AFGR reference panel, the sanger imputation server (https://imputation.sanger.ac.uk/) was used with EAGLE2^61^ for phasing and Positional Burrows-Wheeler transform (PBWT)^62^ for imputation. For the internal reference panel, SHAPEIT4 was used for phasing and Minimac3 was used for imputation. For each imputed SNP, the genotype call was based on the reference panel that yielded the highest imputation quality score. Poorly imputed SNPs (INFO < 0.8) were excluded.

### Combining genotyped and whole-genome sequenced samples

To confirm the accuracy of our imputation approach, for samples that were both genotyped and whole-genome sequenced, genetic concordance between genotypes derived from the two methods was calculated. Across the 27 samples that passed quality control filters on both platforms, an average of 98.995% of SNPs were concordant.

Whole-genome sequenced and genotyped samples (post-imputation) were merged by extracting SNPs common to both methods with overall missingness of less than 10%. The merging was completed using bcftools (v1.15). SNPs that deviate from Hardy-Weinberg equilibrium (P < 5e-5) were excluded, either before or after merging. SNPs with low minor allele frequency (MAF < 0.05) were also excluded after merging. Genetic principal components (PCs) were calculated using PLINK(v1.9) after LD pruning and exclusion of long-range LD regions. Genotype-based principal component analysis (PCA) suggests that there were no systematic batch effects due to platform differences (Supplementary Figure 9A).

To identify TB-DAR samples that are genetic outliers, the TB-DAR samples were merged with 1000 genomes samples and PCA was completed (Supplementary Figure 9B). A K-Nearest Neighbour (KNN) model was trained on the 1000 genomes samples to assign ancestry to four components (European, East Asian, South Asian, African) based on the top 2 PCs. Only one of the TB-DAR samples was not clustered with the African population and was not assigned to the African population by the KNN. This sample was excluded.

For pairs of relatives up to 2nd degree (N = 24), one of the relatives chosen randomly was excluded. In the scenario of trios, the child was excluded. Most likely due to recordkeeping error, a small number of samples (N = 11) was excluded due to discordance between reported and genetically inferred sex.

### Bacterial sequencing

#### Bacterial sequencing

Sputum samples from patients were decontaminated to kill bacteria other than *M*.*tb* and centrifuged to obtain a sputum pellet. The sputum pellet was then inoculated on Löwenstein-Jensen solid media, followed by DNA extraction using the CTAB method and whole-genome sequencing. Culturing was completed at the TB laboratory of the Ifakara Health Institute in Bagamoyo, Tanzania. DNA extraction of bacterial isolates was conducted in Switzerland (before 2017) or in Bagamoyo (after 2017). Sequencing was conducted at the Department of Biosystems Science and Engineering of ETH Zurich, Basel (DBSSE) using Illumina HiSeq 2500 or NovaSeq technology. The bacterial WGS data is published under bioproject PRJEB49562.

#### Bacterial variant calling

The pipeline described by Menardo et al.^63^ was used to process bacterial sequencing reads. In brief, Trimmomatic^64^ (v1.2) was used to trim adapters and low-quality bases. BWA^57^ (v 0.7.13) was used to align reads to the reconstructed ancestral sequence of the MTBC^48^. Duplicate reads were excluded using MarkDuplicates module of Picard^58^ (v2.9.1), and reads with low-quality alignments (>7 mismatches per 100 bp) were excluded using Pysam(v0.9.0).

For variant calling, VarScan^65^ (v2.4.1) mpileup2snp along with SAMtools^66^ (v1.2) mpileup was used. The following VarScan filters were applied: a minimum depth of 7 at a site to make a call (-min-coverage 7), a minimum of 5 supporting reads at a position to call variants (--min-reads2 5), a minimum base quality of 20 at a position to count a read (--min-avg-qual 20), a minimum variant allele frequency threshold of 0.1 (--min-var-freq 0.1), a minimum frequency of 0.9 to call homozygote (--min-freq-for-hom 0.9), and to ignore variants with >90% support on one strand (--strand-filter 1). Non-homozygous mixed calls (called 0|1 or 1|0 heterozygous by varscan) were excluded as they could be due to sequencing errors. Variants in repetitive regions (e.g. PE, PPE, and PGRS genes or phages) were also excluded^67^. A whole-genome FASTA file was then created from the generated VCF file. Samples with a sequencing coverage lower than 15 and those with features suggesting mixed infections (more heterozygous variants than homozygous variants or more than 1000 heterozygous variants or a mix of lineages or sublineages) were excluded from downstream analysis. Lineages and sublineages were defined based on the SNP classification by Steiner et al^68^ and Coll et al^69^ respectively. Genomes were also excluded when multiple sequencing runs of the same DNA samples resulted in different lineage or sublineage assignments.

To annotate variants, SnpEff^70^ (v5.0c) was used. Non-synonymous variants were extracted using the SnpSift module of SnpEff. The resulting VCF files were merged, and a table (0 for absence, 1 for presence, and NA for insufficient coverage) of all amino-acid variants was created using R (v4.4.1).

### Phylogenetic analysis

Using Python v2.7.11 we compiled multiple sequence alignments from the FASTA files and extracted the variable positions with less than or equal to 10% missing data. Phylogenetic trees were constructed using RAxML v8.2.1^71^ with a general time-reversible model of sequence evolution (options -m GTRCAT -v) with a *M*.*canettii* (SAMN00102920) or a L6 genome (SAMEA5366648) as the outgroup. Graphical visualization was created using the ggtree^72^ package in R (v4.4.1).

To understand the prevalence of *M.tb* variants and whether they were under positive selection in a worldwide context, we leveraged the global reference set compiled by Zwyer et al^33^ containing 11,818 genomes using BCFtools v1.15^73^. To formally test for positive selection, PAML v4.9^74^ was run for subsets for each lineage consisting of 300 genomes randomly chosen. As an input we used phylogenetic trees constructed with RAxML v8.2.11 in addition to the gene alignments in order to estimate the branch lengths of the tree based on the M0 codon model in PAML. We then fitted two models to the tree and gene alignment, one modeling nearly neutral (M1a) evolution and the other modeling positive selection (M2a). We calculated p-values based on likelihood ratio tests of the two aforementioned models^75^. We reported the per-site posterior probability based on the Bayes empirical Bayes (BEB) method.

### Genome-to-genome association study

To achieve sufficient statistical power, the G2G study was restricted to common human and *M*.*tb* variants. Variant frequency thresholds were decided based on *a priori* power calculations (Supplementary Figure 1), conducted using genpwr package in R^76^ assuming an additive true and test model. The alpha level was selected based on the genome-wide threshold (5e-8) corrected for the number of independent *M*.*tb* variants tested at each pathogen MAF threshold. On the human side, only variants with MAF > 0.05 were included. This resulted in the inclusion of 6,603,291 human variants. On the bacterial side, only non-synonymous variants with MAF > 0.015 were included. To avoid spurious associations due to stratification, *M*.*tb* variants previously established as phylogenetic lineages or major sublineages^67–69^ were excluded. Lineage markers found in our dataset, defined as variants with high overall frequency (MAF > 0.015) but low frequency (MAF<0.015) within every lineage, were also excluded. After filtering, 1538 *M*.*tb* amino-acid variants were included.

For each *M*.*tb* variant, a case-control GWAS was conducted. Specifically, given *M*.*tb* variant *j*, human variant *i*, and *K* covariates, the model was formulated as:

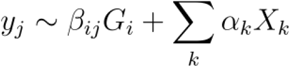

where *y*_*j*_ represents the phenotype vector encoding the presence of *M*.*tb* variant *j, G*_*i*_ represents the genotype dosage vector of human variant *i*, and *X*_*k*_ represents the covariate vector for covariate *k*. Sex and the top three human genetic PCs were included as covariates. The top three PCs were included given that they are among the PCs that capture the most variance and cumulatively capture 29% of the variance in the data (Supplementary Figure 10). The GWAS were conducted using PLINK(v1.9), with a logistic regression model assuming an additive effect. We used Bonferroni correction to adjust for multiple hypothesis testing. To avoid double-counting *M*.*tb* variants that are in perfect correlation (r2=1), the number of independent tests was derived by adding the number of sets of *M*.*tb* variants that are in perfect correlation (N=52) to the number of *M*.*tb* variants that are not in perfect correlation with any other variant (N=436). The genome-wide significance threshold (5e-8) was divided by the number of independent tests (488) to derive a final threshold of 1.02e-10.

### eQTL colocalization study

eQTL summary statistics from 29 studies were downloaded from the eQTL catalogue^77^. Liftover from hg19 to hg38 was completed using the liftOver function of the rtracklayer package (v1.52.1) in R(v4.4.1). Bayesian colocalization analysis implemented by coloc.abf^78^ function of the coloc package(v5.1.0) in R(v4.4.1) was used to conduct colocalization analysis. Default prior probabilities were used. The posterior probabilities for H4 (hypothesis with a single shared casual variant for both traits) were reported. We also checked whether significant eQTLs (p < 5e-8) were identified for our genes of interest (*PRDM15, TIMM21, FBXO15*) in the GENOA cohort^46^, which was based on whole-blood samples from African American individuals. No significant eQTLs were identified, hence we did not include this dataset in the colocalization analyses.

#### M.tb epitope variants and human HLA variation

Experimentally validated *M*.*tb* T-cell epitopes were extracted (October 2022) from the Immune Epitope Database (IEDB). The search query was restricted to the following epitopes: validated by T cell or MHC ligand assays, source organism = *M. tuberculosis* or *M. tuberculosis* H37Rv, and host organism = human. 3,499 epitopes matching the search query were identified. UniProt IDs from query results were mapped to protein names using the UniProt ID mapping tool (https://www.uniprot.org/id-mapping). All common *M*.*tb* amino acid variants (MAF > 0.015) were then mapped to epitopes based on their positions. Human HLA amino acid variants and 4 -digit HLA alleles were imputed using the TopMed Imputation Server (https://imputation.biodatacatalyst.nhlbi.nih.gov/#!) based on the TopMed reference panel^79^.

We tested for associations between *M*.*tb* epitope variants and human HLA amino acid variants or alleles. As with the G2G study, sex and the top three human genetic PCs were included as covariates. The logistic regression-based association model implemented in PLINK(v1.9) was also used but assuming a dominant effect. *M*.*tb* epitope variants in perfect correlation (r2=1) were grouped, resulting in 10 variant sets. We tested the association of 10 *M*.*tb* variant sets against 293 HLA amino acid positions and 103 4-digit HLA alleles. A Bonferroni corrected threshold of 1.3e-5 was thus applied, based on an alpha level of 0.05 and the 3960 tests we conducted.

### Pathway-to-pathway association study

Human gene regulatory pathways were based on the 6449 curated gene-sets from Human Molecular Signatures Database (MSigDB, v7.0)^80^. The curated gene sets were generated from either gene expression profiles of perturbation experiments or from canonical pathway databases including Reactome, KEGG and others. For each GWAS conducted in the G2G study, gene scores were constructed by mapping variants to genes by proximity using PASCAL^81^. Given that variants in close proximity are likely to be in strong LD, gene scores of neighboring genes are often correlated and can be part of the same pathway. To address this, PASCAL constructs a fusion score based on LD patterns. The 1000 Genomes African population was used as the reference LD database for this.

*M*.*tb* gene regulatory pathways from two data sources were obtained. 547 pathways were derived from a *M*.*tb* regulatory network constructed using gene expression profiles of perturbation experiments(http://networks.systemsbiology.net/mtb/)^82^. 192 pathways were derived from the stringDB^83^ database using a score-cutoff of 400 and recursive MCL clustering with inflation parameter optimized to obtain the maximum number of clusters between the size of 5 - 50. *M*.*tb* variants were directly mapped to *M*.*tb* genes based on position.

To identify the enrichment of edges between pairs of human-bacterial pathways, p-values between human and *M*.*tb* genes were binarized based on False Discovery Rate (FDR) < 0.15. A density measure was calculated for each pair of human and *M*.*tb* pathways. Given a human pathway with gene-set 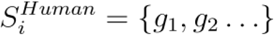, a *M*.*tb* pathway with gene-set 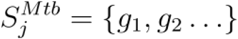, and a gene-gene binary association matrix X, the density was defined as:

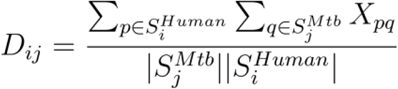

where *D*_*ij*_ represents the density between human pathway *i* and *M.tb* pathway *j*.

To calculate an empirical p-value, gene-pathway memberships were permuted 1000 times while preserving the size of the pathways. The distribution of density values obtained from the permutations was used to obtain a null distribution.

## Data Availability

The bacterial WGS data has been published under bioproject PRJEB49562. Human genotyping and WGS data are deposited on the European Genome-phenome Archive (EGA) under EGAS00001007216 and EGAS00001005850 respectively.

## Acknowledgement

This work was supported by the Swiss National Science Foundation (Grants: CRSII5-177163 and 310030-188888) and the European Research Council (Grant: 883582). We thank E. Ristorcelli (CHUV, Lausanne, Switzerland) for sample preparation and DNA extraction; we thank K. Harshman, I. Bartha, C. Howald, and D. Lamparter (Health 2030 Genome Center, Geneva, Switzerland) for human WGS support; we thank M. Docquier (iGE3 Genomics Platform, Geneva, Switzerland) for human genotyping support.

## Data Availability

Summary statistics and software code have been published on github: https://github.com/zmx21/G2G-TB. The bacterial WGS data has been published under bioproject PRJEB49562. Human genotyping and WGS data are deposited on the European Genome-phenome Archive (EGA) under EGAS00001007216 and EGAS00001005850 respectively.

