## Supplementary Materials for "Genome-to-genome analysis reveals associations between human and mycobacterial genetic variation in tuberculosis patients from Tanzania"

### Supplementary Tables and Figures

#### Supplementary Table 1

Association between Rv2348c I101M and host variant (rs12151990), covariates, and patient characteristics based on logistic regression. The minor allele of rs12151990 (T) was encoded as the effect allele.

| | log(OR) | SD | P |
|---|---|---|---|
| rs12151990 | 1.73 | 0.26 | **4.67e-11** |
| **Covariates** | | | |
| Sex | -7.7e-3 | 9.2e-3 | 0.40 |
| Human PC1 | -0.03 | 0.16 | 0.86 |
| Human PC2 | 0.14 | 0.16 | 0.40 |
| Human PC3 | -0.02 | 0.16 | 0.92 |
| **Patient Characteristics** | | | |
| Age | -3.8e-4 | 4.2e-3 | 0.93 |
| BMI | 2.3e-3 | 4.2e-3 | 0.59 |
| Smoker | 3.6e-3 | 9.9e-3 | 0.72 |
| HIV Positive | 6.9e-3 | 0.01 | 0.55 |

Supplementary Table 2

Association between FixA T67M and host variant (rs75769176), covariates, or patient characteristics based on logistic regression. The minor allele of rs75769176 (G) was encoded as the effect allele.

| | log(OR) | SD | p |
|---|---|---|---|
| rs75769176 | 1.9 | 0.29 | **6.28e-11** |
| **Covariates** | | | |
| Sex | 5.8e-4 | 0.01 | 0.95 |
| Human PC1 | 0.27 | 0.18 | 0.14 |
| Human PC2 | -0.56 | 0.18 | **2.3e-3** |
| Human PC3 | -0.25 | 0.18 | 0.165 |
| **Patient Characteristics** | | | |
| Age | -2.8e-3 | 4.7e-3 | 0.55 |
| BMI | 8.1e-3 | 4.7e-3 | 0.09 |
| Smoker | -3.1e-3 | 0.01 | 0.78 |
| HIV Positive | -0.01 | 0.01 | 0.41 |

Supplementary Table 3

**Association between clinical measures and genome-to-genome (G2G) associated variants.** Ordinal logistic regression was used for TB score and linear regression was used for X-ray score and GeneXpert Ct-value. Estimates correspond to beta for linear regression and log odds for ordinal regression. Covariates were selected using bidirectional stepwise selection.

| Dependent Variable | Covariates | Independent Variable | p | Estimate (SE) |
|---|---|---|---|---|
| TB score | Cough_Duration + Sex + Smoking + HIV Status | rs12151990 | 0.76 | 0.04 (0.13) |
| | | Rv2348c I101M | 0.78 | 0.12 (0.41) |
| | | **rs75769176** | **0.04** | **-0.34 (0.17)** |
| | | FixA T67M | 0.13 | -0.58 (0.38) |
| X-ray score | Age + Sex + HIV Status | rs12151990 | 0.22 | 0.13 (0.10) |
| | | Rv2348c I101M | 0.21 | 0.39 (0.31) |
| | | rs75769176 | 0.87 | -0.02 (0.12) |
| | | FixA T67M | 0.76 | 0.09 (0.30) |
| Ct value | HIV Status + Cough Duration + Smoking | rs12151990 | 0.86 | 0.02 (0.10) |
| | | Rv2348c I101M | 0.64 | -0.19 (0.40) |
| | | rs75769176 | 0.38 | 0.12 (0.13) |
| | | FixA T67M | 0.09 | 0.48 (0.29) |

Supplementary Table 4

Human variants previously identified to be associated with susceptibility for developing active TB. The top G2G association for each human variant is displayed. If the query variant was not common within our cohort, proxy variant indicates a human variant within ±5kb that displays the strongest association with a *M.tb* variant.

| Study | Human Variant (Gene) | Proxy Variant | *M.tb* Variant | P | OR |
|---|---|---|---|---|---|
| **Associations with TB susceptibility** | | | | | |
| Thye et al, 2012[14]<br>Chimusa et al, 2014[15] | rs2057178 (*WT1*) | | Rv0178:p.Asp150Glu | 1.8e-2 | 0.6 |
| Thye et al, 2010[16] | rs4331426 | | hsdM:p.Lys483Glu | 5.5e-2 | 1.7 |
| Curtis et al, 2015[17] | rs10956514 (*ASAP1*) | | eccE1:p.Phe113fs | 4.4e-2 | 1.4 |
| | rs4733781 (ASAP1) | | pyrH:p.Ala247Val | 3.3e-3 | 2.4 |
| Zheng et al, 2018[18] | rs12437118 (*ESRRB*) | | Rv2078:p.Gly74Asp | 1.8e-3 | 0.4 |
| | rs6114027 (*TGM6*) | | fixA:p.Thr67Met | 4.3e-4 | 0.3 |
| Luo et al, 2019[19] | rs73226617 (*ATP1B3*) | rs4683619 | fixA:p.Thr67Met | 1.8e-3 | 0.4 |
| Kerner et al, 2019[20] | rs34536443 (*TYK2*) | rs203999 | PE35:p.Met4fs | 1.7e-3 | 0.4 |
| Quistrebert et al, 2021[21] | rs17155120 (C10orf90/ADAM21) | | Rv2425c:p.Met289Ile | 7.6e-3 | 1.6 |
| Qi et al, 2017[22] | rs4240897 (*MFN2*) | | Rv2963:p.Gly165fs | 9.5e-5 | 1.4 |
| Sobota et al, 2016[23] | rs4921437 (*IL12B*) | | ltp1:p.Ser6fs | 8.2e-4 | 2.4 |

Supplementary Table 5

Human variants previously identified to be associated with lineages or clades of *M.tb*. The top G2G association for each human variant is displayed. If the query variant was not common within our cohort, proxy variant indicates a human variant within ±5kb that displays the strongest association with a *M.tb* variant.

| Study | Human Variant (Gene) | Proxy Variant | *M.tb* Variant | P | OR |
|---|---|---|---|---|---|
| **Associations with *M.tb* Lineage / Clade** | | | | | |
| McHenry et al, 2020[36] | rs17235409 (*SLC11A1*) | rs146731249 | hsdM:p.Lys483Glu | 5.0e-6 | 4.8 |
| McHenry et al, 2021[37] | rs114945555 (*PPIAP22*) | | Rv3916c:p.Leu221Val | 6.6e-2 | 2.5 |
| Luo et al, 2022[34] | rs3130660 (*FLOT1*) | rs113903830 | Rv2179c:p.Glu30Lys | 6.1e-3 | 1.7 |
| Müller et al, 2021[38] | rs529920 | | Rv0574c:p.Tyr45His | 1.3e-2 | 2.0 |
| | rs41472447 (*PDZRN4*) | rs36070978 | Rv0104:p.Ile56Ser | 3.7e-3 | 3.4 |
| Phelan et al, 2023[35] | rs267951 (*DAP*) | | helZ:p.Ile727Leu | 1.4e-2 | 0.4 |
| | rs74875032 | rs4900336 | Rv2949c:p.Trp189Leu | 2.2e-4 | 2.4 |
| | rs60284130 (*MFAP2*) | rs56265117 | Rv0435c:p.Ala61Thr | 1.1e-3 | 0.4 |
| | rs142600697 (*FSTL5*) | rs61381755 | Rv3847:p.Ile170Thr | 1.1e-5 | 3.9 |
| | rs1118438 | | Rv0377:p.Arg302Pro | 2.9e-2 | 0.8 |
| | rs558237 (*RIMS3*) | | Rv2134c:p.Asp192Ala | 3.0e-2 | 0.6 |
| | rs59441182 | | Rv2026c:p.Pro102Thr | 2.1e-2 | 0.5 |
| | rs4563899 (*CSGALNACT*) | | Rv0839:p.Ala205Thr | 2.4e-3 | 0.4 |

Supplementary Table 6

Experimentally validated epitopes (according to IEDB) that each *M.tb* variant maps to (with location of variant indicated in square brackets within epitope sequence). If an epitope has been experimentally validated to be restricted by an HLA-DRB1 allele, the respective alleles are indicated.

| ***M.tb*<br>protein** | ***M.tb*<br>variant** | ***M.tb*<br>T cell epitope** | **IEDB experiment -<br>human HLA-DRB1 restriction** |
|---|---|---|---|
| EspK | L39W | DTFYDRAQEYSQV[L>W]Q | N/A |
| EsxB | E68K | AAGTAAQAAVVRFQEAANKQKQ[E>K]LD,<br>AANKQKQ[E>K]LDEISTN,<br>AANKQKQ[E>K]LDEISTNIRQAG,<br>AANKQKQ[E>K]LDEISTNIRQAGVQYSR,<br>AAVVRFQEAANKQKQ[E>K]L,<br>AQAAVVRFQEAANKQKQ[E>K]LD,<br>[E>K]LDEISTNIRQAGVQYSR,<br>KQ[E>K]LDEISTNIRQAG,<br>RFQEAANKQKQ[E>K]LDE,<br>VRFQEAANKQKQ[E>K]LD,<br>VVRFQEAANKQKQ[E>K]L,<br>AVVRFQEAANKQKQ[E>K],<br>[E>K]LDEISTNIRQAGVQ,<br>FQEAANKQKQ[E>K]LDEI,<br>Q[E>K]LDEISTNIRQAGV | N/A<br>HLA-DRB1*04:05<br>N/A<br>N/A<br>N/A<br>N/A<br>N/A<br>HLA-DRB1*03:01,04:01,08:02,11:01,13:02<br>N/A<br>HLA-DRB1*04:01,04:04,04:05,09:01,11:01<br>N/A<br>N/A<br>N/A<br>N/A<br>N/A |
| Mpt70 | A21T | GLAAL[A>T]VAVSPPAAAGDLVGPGCAE,<br>MKVKNTIAATSFAAAGLAAL[A>T]VAVS,<br>SFAAAGLAAL[A>T]VAVSPPAA | N/A<br>N/A<br>N/A |
| RimJ | R72L | G[R>L]MLPYVIEL | N/A |

Supplementary Table 7

**Filters recommended by Illumina to exclude poor quality probes.** Minor allele frequency (MAF) based filters were not applied to X-Chr probes.
Adapted from:
https://www.illumina.com/Documents/products/technotes/technote_infinium_genotyping_data_analysis.pdf

| **Metric(s)** | **Threshold for exclusion** |
|---|---|
| Rep Errors | > 2 |
| PPC Errors | > 2 |
| Cluster Sep | ≤ 0.3 |
| AA R Mean | ≤ 0.2 |
| AB R Mean | ≤ 0.2 |
| BB R Mean | ≤ 0.2 |
| 10%_GC_Score | ≤ 0.3 |
| Hex Excess | > 0.2 |
| A/B_Freq | ≥ 0.4 |
| AB T Mean | (< 0.2) or (> 0.8) |
| A/A_Freq = 1 | |
| AA T Mean | > 0.3 |
| AA T Dev | > 0.06 |
| B/B_Freq = 1 | |
| BB T Mean | < 0.7 |
| BB T Dev | > 0.06 |
| A/A_Freq or B/B_Freq = 0 | |
| AB T Dev | > 0.05 |
| A/B_Freq = 0 | |
| MAF | > 0 |
| 0.998 > Call_Freq > 0.990 | |
| MAF | < 0.05 |

Supplementary Figure 1

Statistical power calculation assuming an alpha level that accounts for multiple testing (5e-8 / Number of independent *M.tb* variants at specific MAF threshold). An additive test and true model was assumed. Statistical power is shown for each variant effect size measured by odds ratio (OR), human variant minor-allele frequency (MAF), and *M.tb* minor-allele frequency (*M.tb* MAF). A *M.tb* MAF threshold of 0.015 was chosen, given that for *M.tb* variants with lower frequency there is very low statistical power to detect associations with common host variants (MAF > 0.05) even under the assumption of a very high effect size (OR = 25).

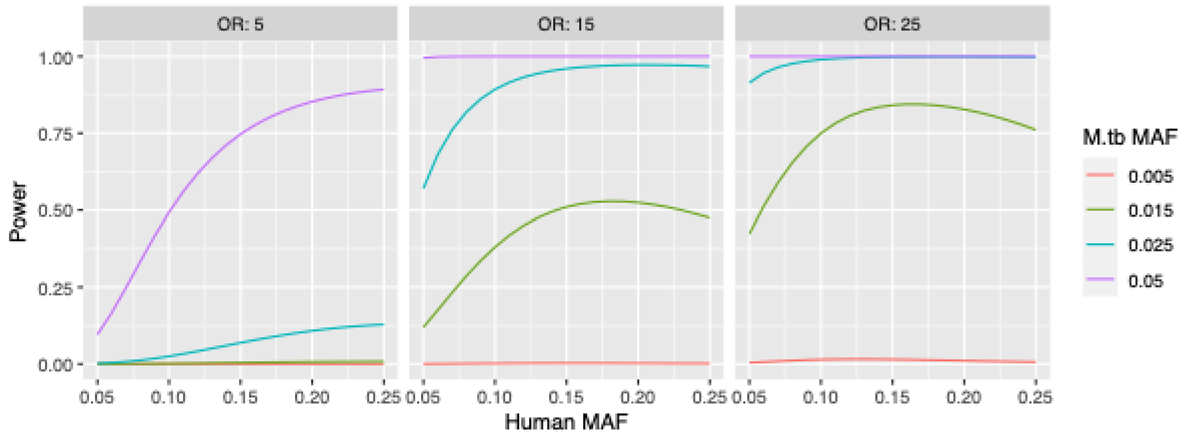

Supplementary Figure 2

Genomic inflation factor (λ) for all GWAS conducted as part of the genome-to-genome (G2G) study.

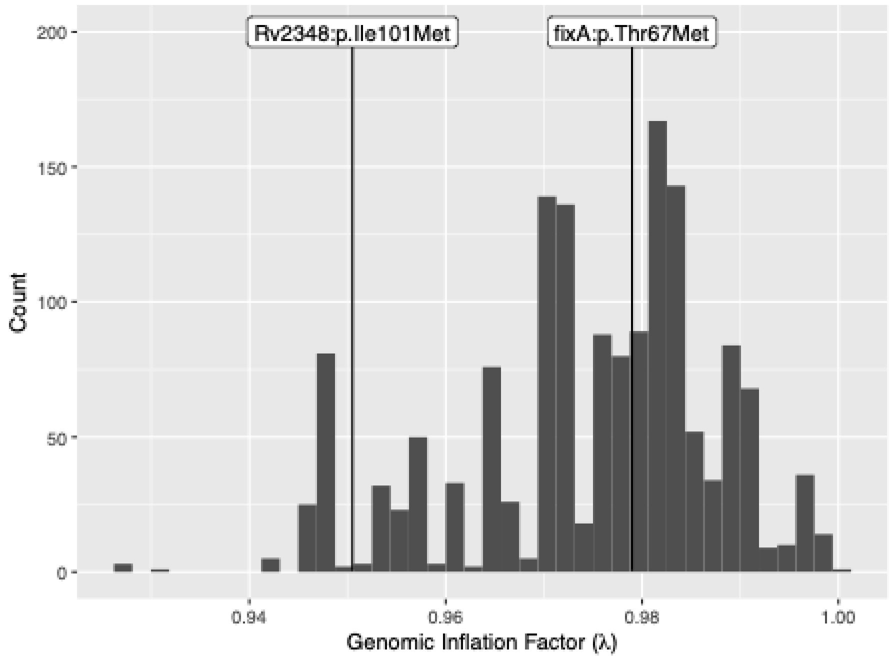

Supplementary Figure 3

*M.tb* variants that are in perfect correlation were grouped into variant-sets. Variant-sets of size 1 indicate independent *M.tb* variants (N = 436). Variant-sets of size greater than one (N=52) indicate groups of perfectly correlated variants. 1102 variants belong to such variant-sets.

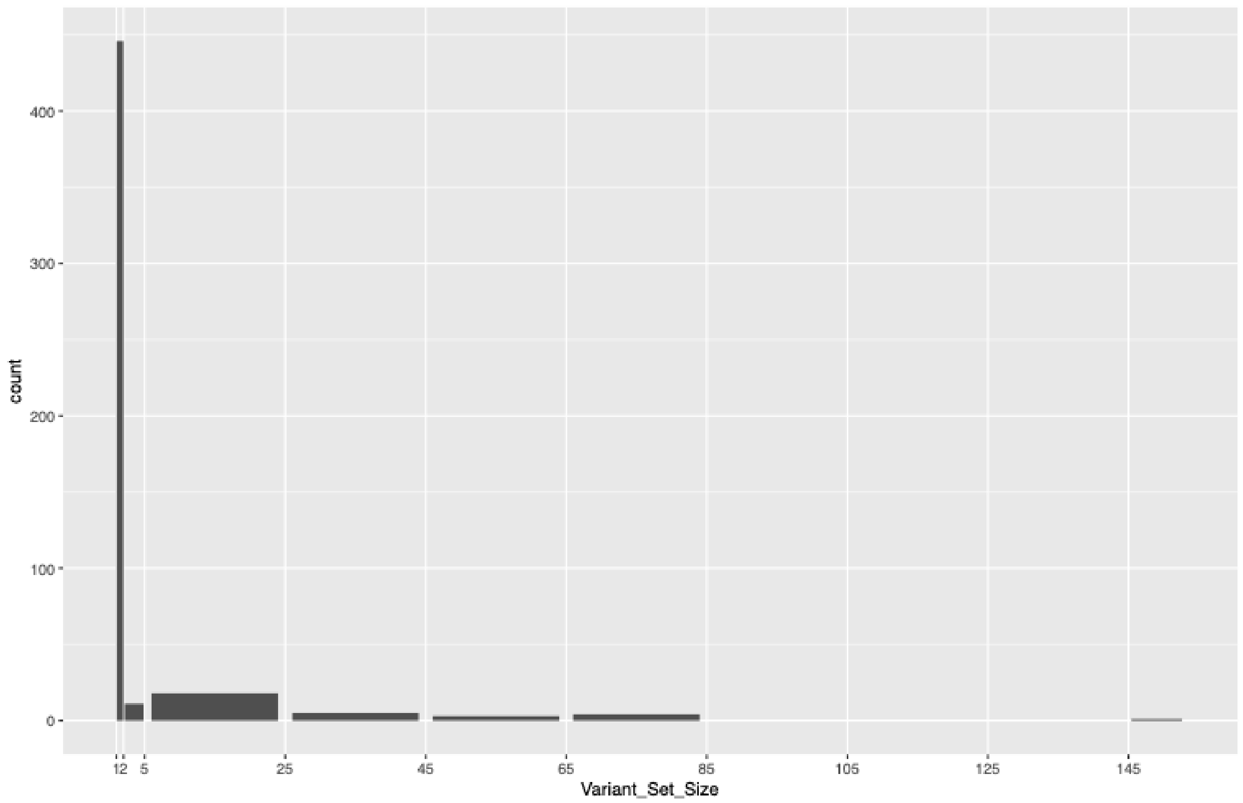

Supplementary Figure 4

Each dot represents a genome-to-genome (G2G) association between a human and *M.tb* variant, with colors indicating p-value. The two significant associations are circled.

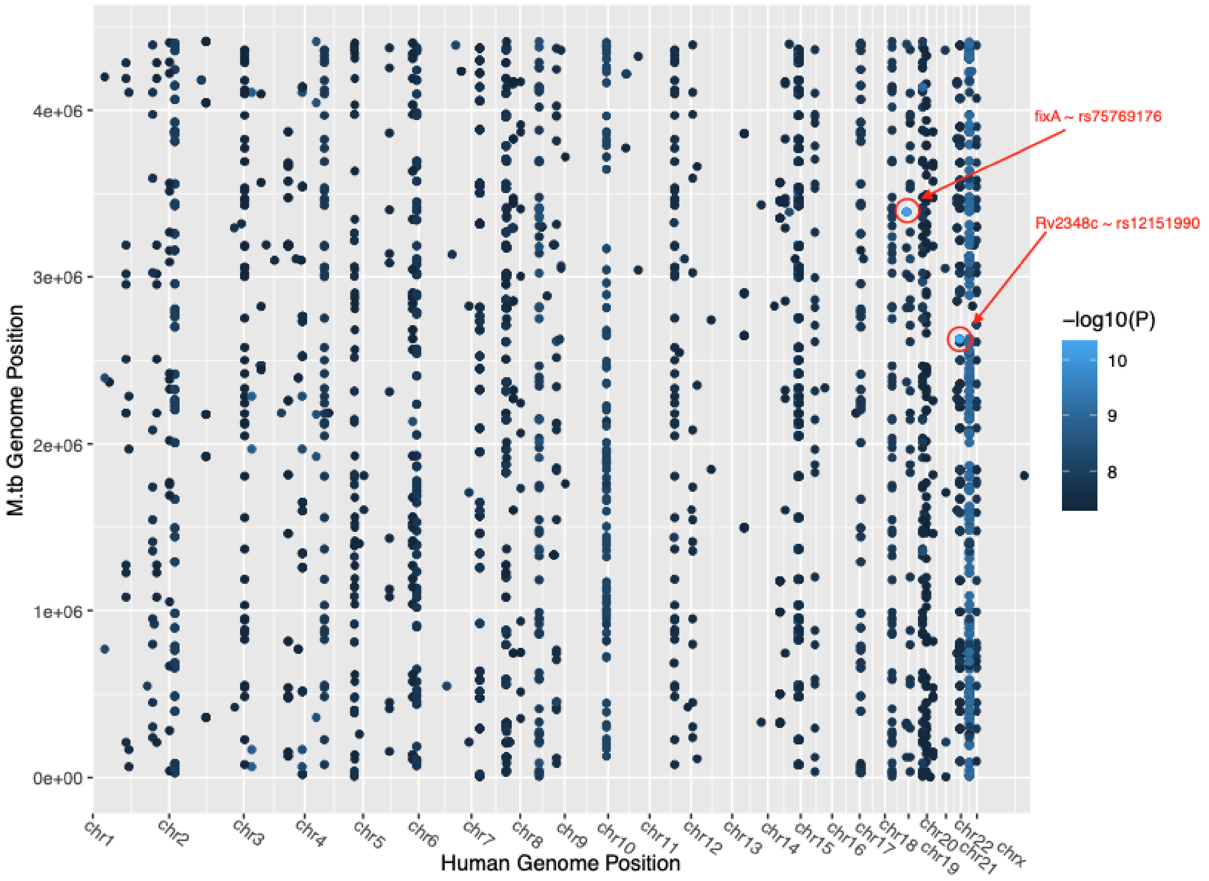

Supplementary Figure 5

Locus zoom plot of **A)** human genetic loci of association between Rv2348c I101M and rs12151990 **B)** human genetic loci of association between FixA T67M and rs75769176

**A)**

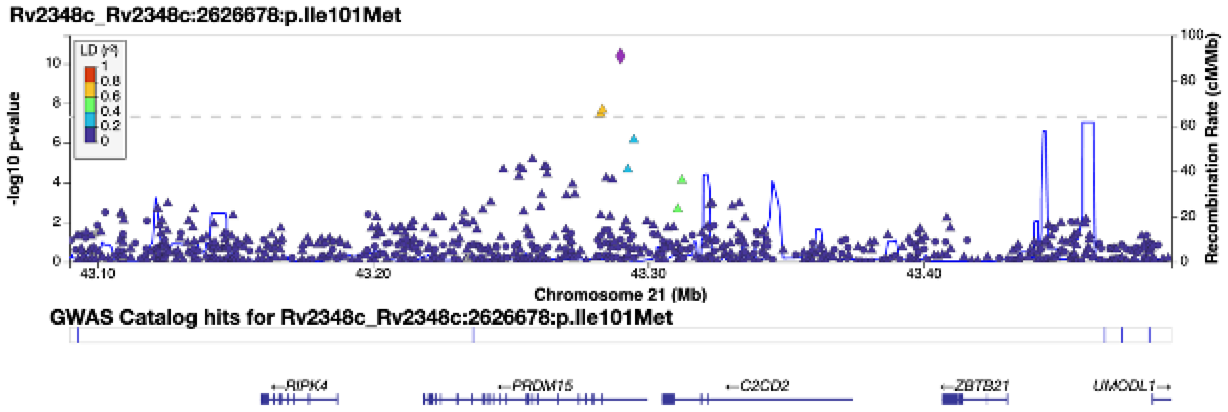


**B)**

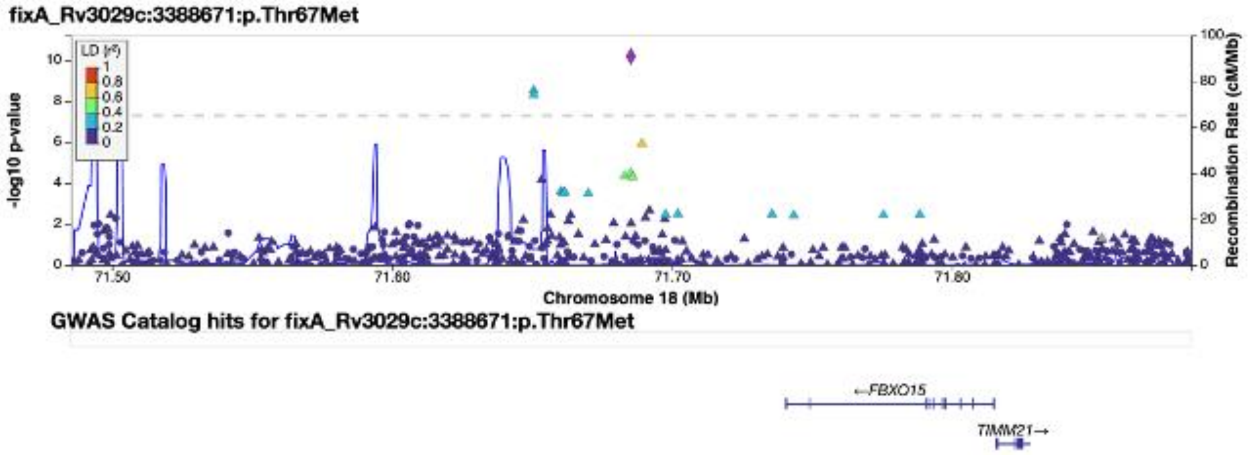

#### Supplementary Figure 6

Posterior probability of shared causal variant based colocalization of GWAS and eQTL signals across different tissues. **A)** between rs12151990 and eQTLs of *PRDM15* **B)** between rs75769176 and eQTLs of *FBXO15* **C)** between rs75769176 and eQTLs of *TIMM21*

**A)**

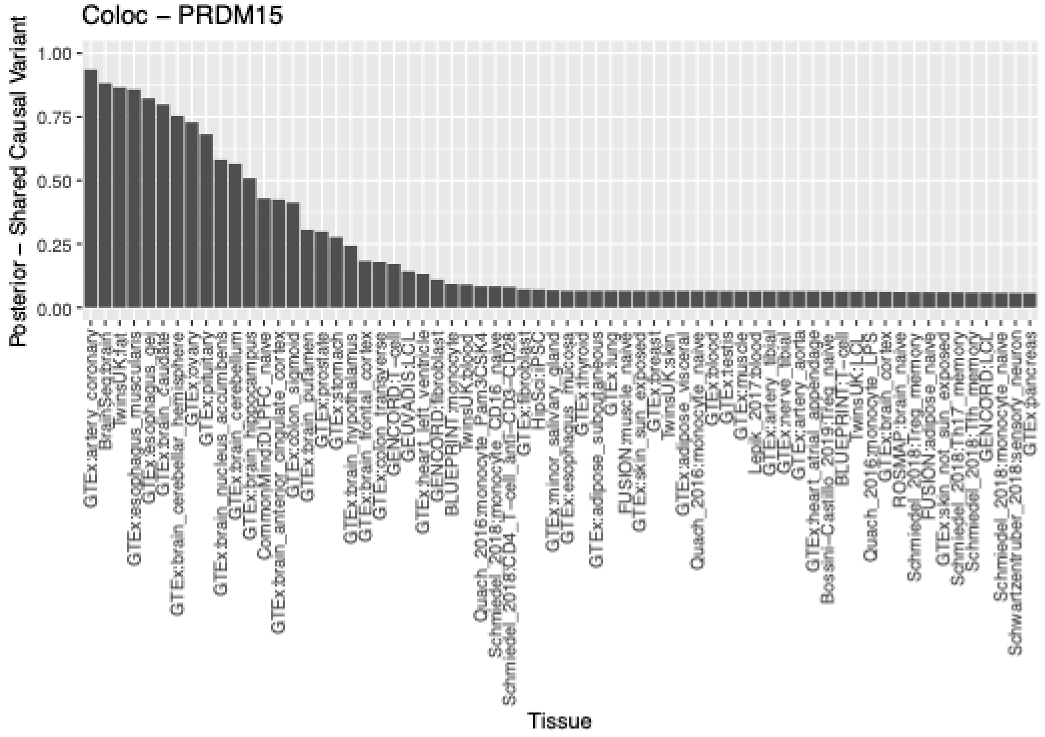

**B)**

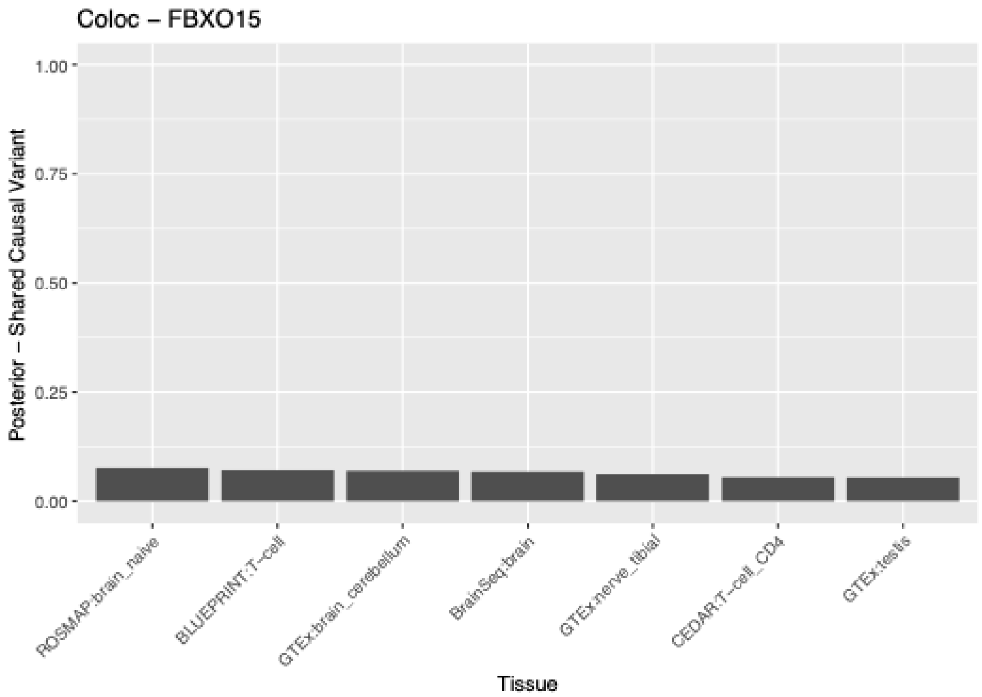

Coloc – FBXO15
Posterior – Shared Causal Variant
1.00
0.75
0.50
0.25
0.00
ROSMAP:brain_naive
BLUEPRINT:T−cell
GTEx:brain_cerebellum
BrainSeq:brain
GTEx:nerve_tibial
CEDAR:T−cell_CD4
GTEx:testis
Tissue


**C)**

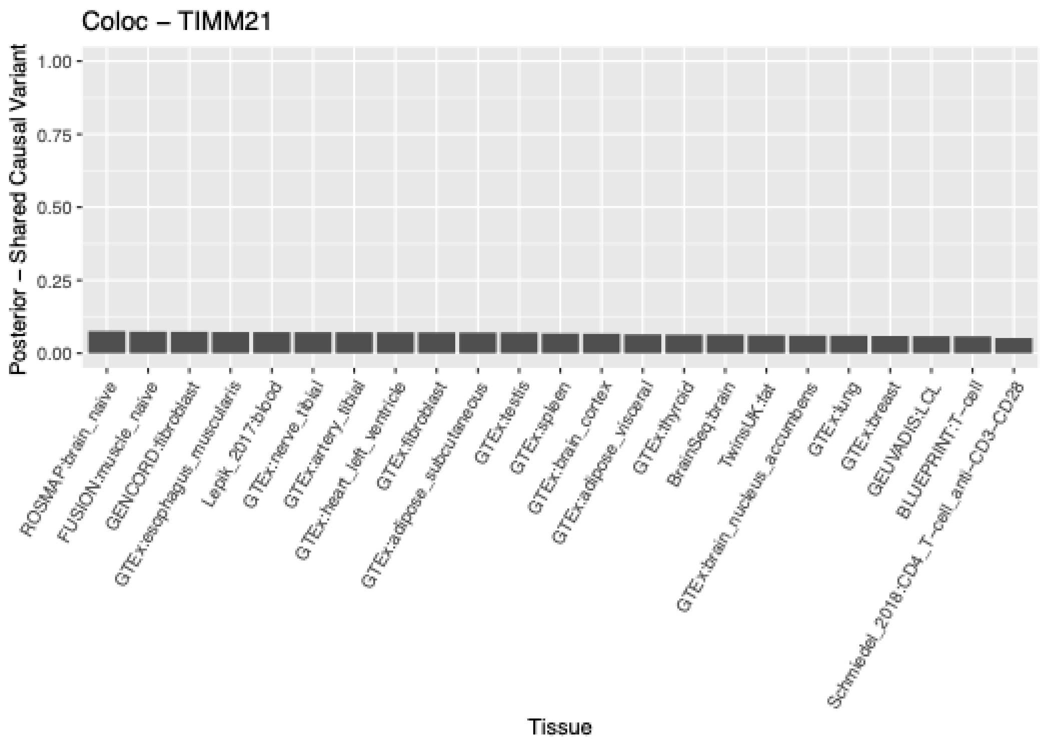

Coloc – TIMM21
Posterior – Shared Causal Variant
1.00
0.75
0.50
0.25
0.00
ROSMAP:brain_naive
FUSION:muscle_naive
GENCORD:fibroblast
GTEx:esophagus_muscularis
Lepik_2017:blood
GTEx:nerve_tibial
GTEx:artery_tibial
GTEx:heart_left_ventricle
GTEx:fibroblast
GTEx:adipose_subcutaneous
GTEx:testis
GTEx:spleen
GTEx:brain_cortex
GTEx:adipose_visceral
GTEx:thyroid
BrainSeq:brain
TwinsUK:fat
GTEx:brain_nucleus_accumbens
GTEx:lung
GTEx:breast
GEUVADIS:LCL
BLUEPRINT:T−cell
Schmiedel_2018:CD4_T−cell_anti−CD3−CD28
Tissue

Supplementary Figure 7

Distribution of module-module density values based on permuted dataset. P-value indicates significance of a density of 0.11, corresponding to the pair of human and *M.tb* modules with the highest density.

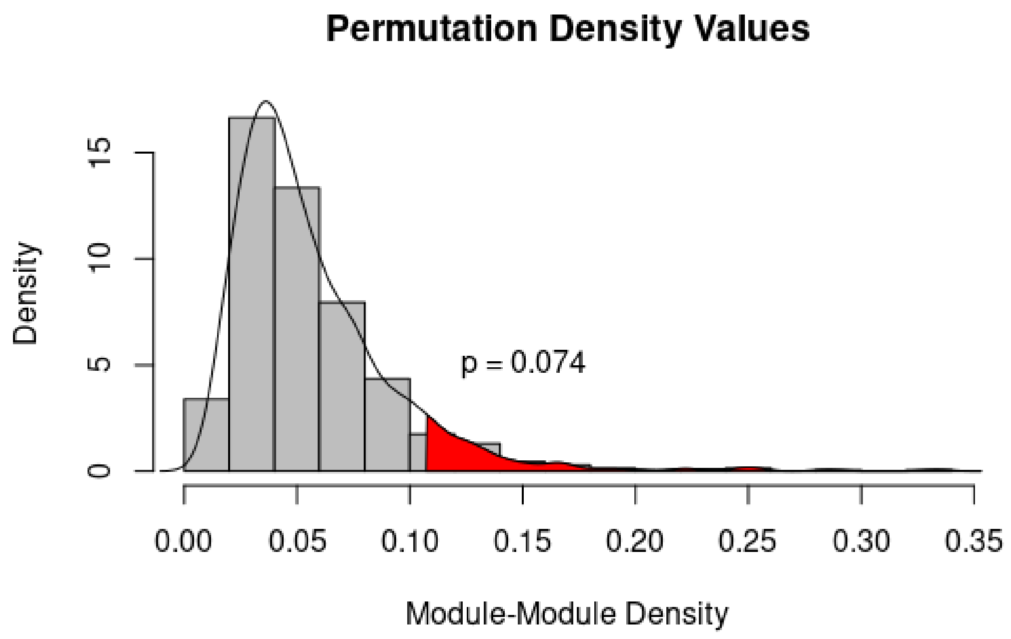

Supplementary Figure 8

Location of *M.tb* epitope variants based on the H37Rv reference genome. Perfectly correlated variants are grouped into SNP sets and labeled with different colors.

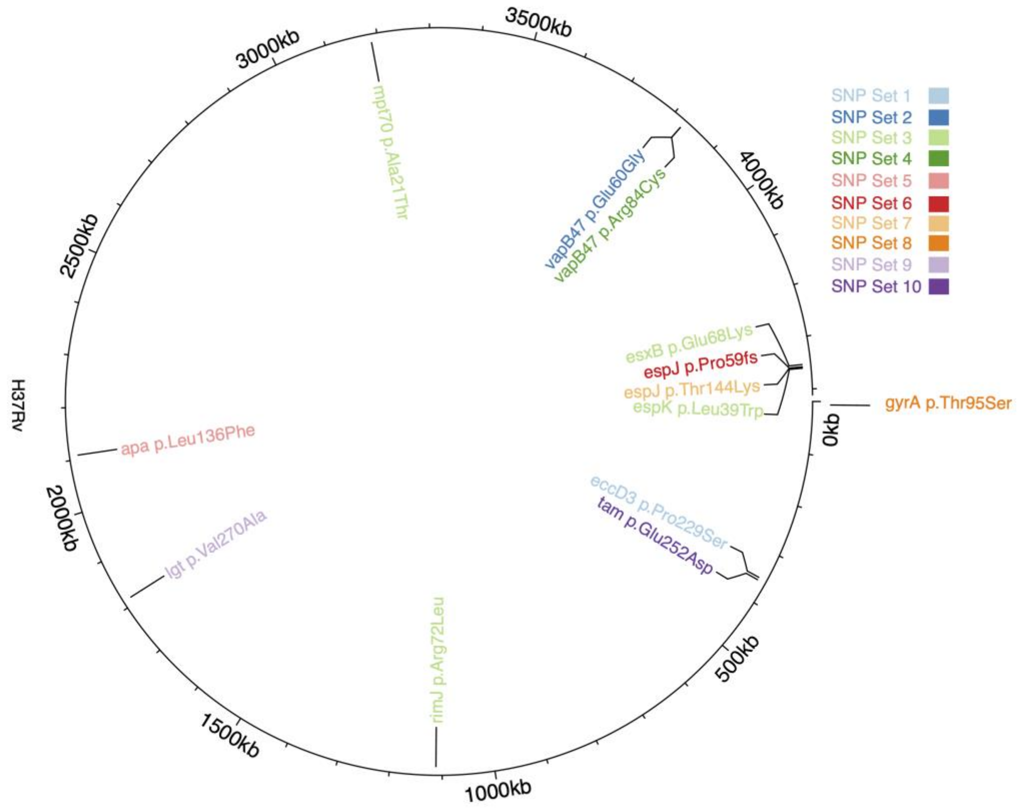

Supplementary Figure 9

**A)**

Top two human genetic principal components (PCs) based on a merged dataset of our cohort (TBDAR) and 1000 Genome superpopulations. A single outlier within our cohort that was not assigned to the African population is labeled.

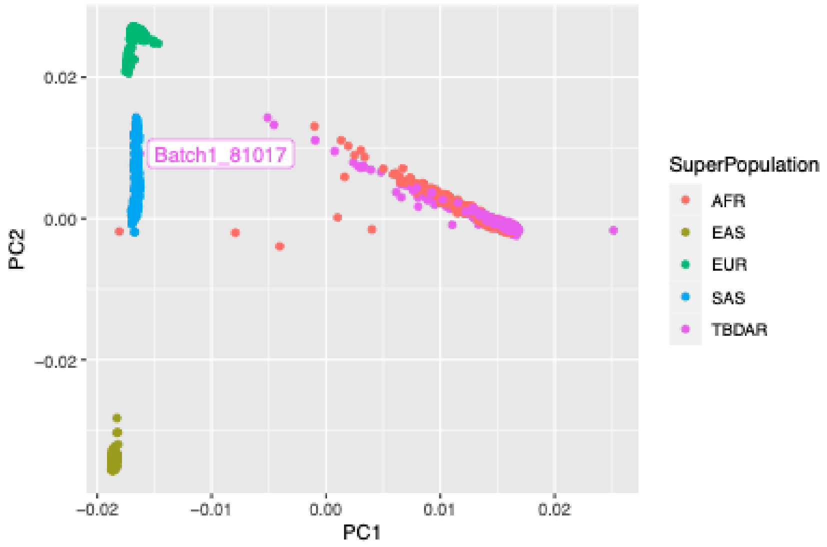


**B)**

Top two human genetic principal components (PCs) of our cohort. Variance explained is indicated in brackets. Colors indicate whether the sample was genotyped and imputed or whole-genome sequenced.

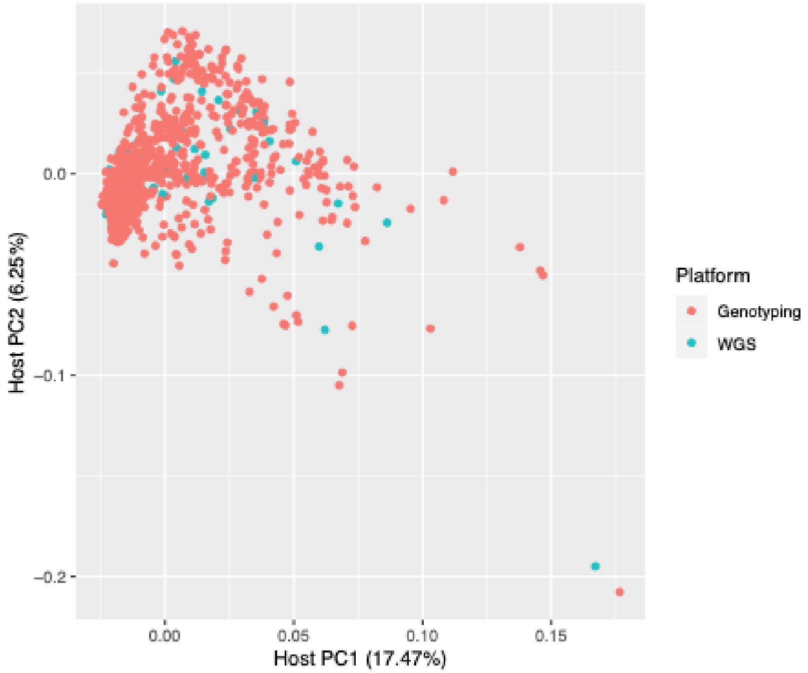

Supplementary Figure 10

Variance explained by each human genetic principal component (PC).

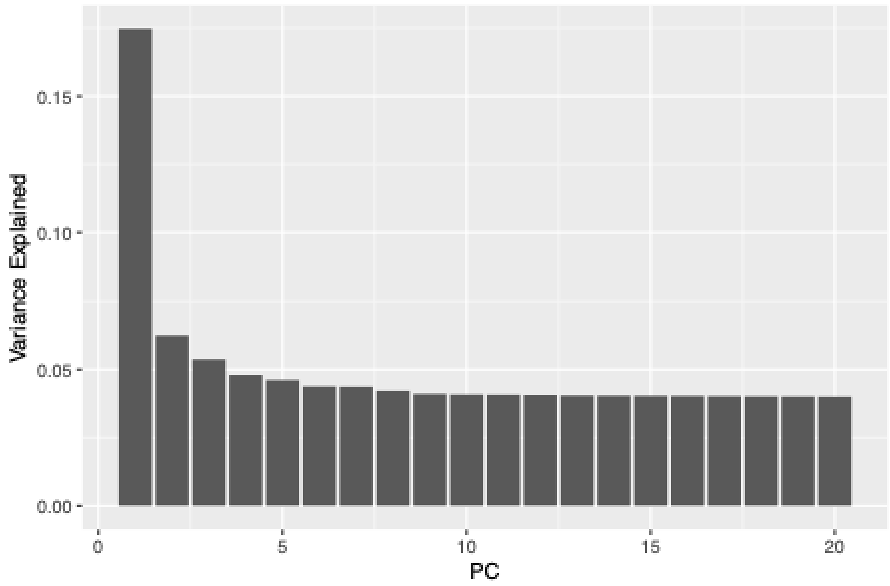
